# DNA Methylation in Cocaine Use Disorder – An Epigenome-wide Approach in the Human Prefrontal Cortex

**DOI:** 10.1101/2022.11.05.22281974

**Authors:** Eric Poisel, Lea Zillich, Fabian Streit, Josef Frank, Marion M Friske, Jerome C Foo, Naguib Mechawar, Gustavo Turecki, Anita C Hansson, Markus M Nöthen, Marcella Rietschel, Rainer Spanagel, Stephanie H Witt

## Abstract

Cocaine use disorder (CUD) is characterized by a loss of control over drug intake and is associated with structural, functional, and molecular alterations in the brain. At the molecular level, epigenetic alterations are hypothesized to contribute to the higher-level functional and structural brain changes as observed in CUD. Most evidence of cocaine-associated epigenetic changes comes from animal studies while only a few studies have been performed using human tissue. We investigated epigenome-wide DNA methylation signatures of CUD in human postmortem brain tissue of Brodmann Area 9 (BA9). A total of N = 42 BA9 brain samples were obtained from N = 21 individuals with CUD and N = 21 individuals without a CUD diagnosis. We performed an epigenome-wide association study and analyzed CUD-associated differentially methylated regions (DMRs). To assess the functional role of CUD-associated differential methylation, we performed Gene Ontology enrichment analyses and characterized co-methylation networks using a weighted correlation network analysis. We further investigated epigenetic age in CUD using epigenetic clocks for the assessment of biological age. While no CpG site was associated with CUD at epigenome-wide significance in BA9, we detected a total of 20 CUD-associated DMRs. After annotation of DMRs to genes, we identified *NPFFR2* and *KALRN* for which a previous role in the behavioral response to cocaine in rodents is known. Three of the four identified CUD-associated co-methylation modules were functionally related to neurotransmission and neuroplasticity. Protein-protein interaction networks derived from module hub genes revealed several addiction-related genes as highly connected nodes such as *CACNA1C, NR3C1*, and *JUN*. In BA9, we observed a trend toward epigenetic age acceleration in individuals with CUD remaining stable even after adjustment for covariates. Results from our study highlight that CUD is associated with epigenome-wide differences in DNA methylation levels in BA9 particularly related to synaptic signaling and neuroplasticity. This supports findings from previous studies that report on the strong impact of cocaine on neurocircuits in the human prefrontal cortex. Further studies are needed to follow up on the role of epigenetic alterations in CUD focusing on the integration of epigenetic signatures with transcriptomic and proteomic data.

## Introduction

Cocaine is one of the most frequently consumed psychostimulants in the world with around 21 million individuals having used cocaine in 2020 [1]. During the last decade, a stable increase in cocaine consumption and cocaine-associated hospitalization rates has been observed [1, 2]. While a substantial fraction of individuals retain control over cocaine intake, up to 21% of regular cocaine users develop cocaine dependence characterized by repeated cycles of binge intoxication, withdrawal, and compulsive cocaine-seeking [3]. Chronic use of cocaine, especially crack cocaine, increases the risk for the development of cardiovascular, cerebrovascular, and respiratory diseases [4]. Also, it is associated with substance abuse of other drugs such as alcohol, cannabis, and opioids [5]. Thus, chronic cocaine consumption as observed in cocaine use disorder (CUD) enhances the overall disease burden, at the individual level significantly reduces life quality, and thereby contributes to an increased risk for premature death. Treatment of CUD is currently only symptomatic, and no FDA-approved drug exists [6]. It is thus of crucial importance to better understand the pathophysiology of CUD to provide a basis for the development of new therapeutic approaches.

Substance use disorders (SUDs) including CUD are understood as brain disorders with profound alterations at the structural, functional, and molecular levels in the human brain [7-9]. Drug-induced effects on epigenetics are hypothesized to contribute to structural and functional changes in the brain via the establishment of altered transcriptional programs (for Review see [10-12]). Thus, profiling of epigenetic alterations contributes to a better understanding of molecular mechanisms underlying the neuroplastic changes associated with CUD. DNA methylation (DNAm) represents the most prominent epigenetic mechanism involved in gene expression regulation and DNAm changes in the brain following acute and chronic cocaine exposure are well described [13]. So far, the effects of cocaine on DNAm were most extensively studied in the rodent brain, which has led to the identification of cocaine-associated differentially methylated genes. Interestingly, several of these genes were previously related to the neurobiology of addiction, such as *Bdnf* [14] and *Pp1c* [15] as well as the immediate early genes *c-Fos* [16] and *fosB [15]*. Cocaine was also shown to affect methylation and expression levels of DNA methyltransferases (Dnmt1, Dnmt3a) [16-19] and Tet demethylases [20] thereby directly altering DNAm homeostasis.

In contrast, much less is known about cocaine-associated DNAm changes in humans. Due to an easy and minimally invasive sampling procedure, epigenome-wide association studies (EWASs) of different cocaine use phenotypes have been performed in human blood samples [21, 22]. These studies revealed an enrichment of cocaine-associated differential methylation within biological pathways involved in synaptic transmission, neurogenesis, cell division, and chromatin organization. However, as DNAm levels are known to be tissue-specific, the predictive value of differential methylation in blood for methylation signatures of CUD in the brain might be limited. Thus, profiling DNAm levels in brain tissue is of critical importance. So far, two epigenome-wide characterizations in CUD have been performed in human postmortem brain tissue from the caudate nucleus (CN) [23] and the nucleus accumbens (NAc) [24]. By analyzing postmortem tissue of the CN from N = 25 CUD cases and N = 25 controls using a reduced representation bisulfite sequencing approach, a total of 173 differentially methylated regions (DMRs) were found that were significantly associated with CUD status. The *IRX2* gene was among the top findings of CUD-associated differential methylation in the CN. Using a similar approach in the NAc, the key finding was a hypermethylated region within the tyrosine hydroxylase (*TH*) gene encoding an enzyme involved in dopamine metabolism. Interestingly, the identified DMR in *TH* was shared between the CN and the NAc. Both studies provide some first mechanistic insights into the biological relevance of CUD-associated DNAm changes in the human brain.

While epigenome-wide alterations associated with CUD have been studied in the human striatum, they have not been reported for cortical brain regions so far. The prefrontal cortex (PFC) has a key role in the addiction cycle due to its involvement in inhibitory control and response to drug-related cues both depicting dysregulated cognitive processes in SUDs [25]. In CUD, resting-state connectivity alterations have been found in frontostriatal circuits representing functional connections between the PFC and the striatum [26]. As DNAm alterations in the PFC have been described for the Brodmann Area 9 (BA9) subregion in other SUDs such as alcohol use disorder (AUD) [27, 28] and opioid use disorder (OUD) [29, 30], BA9 depicts a promising brain region for the epigenome-wide profiling of DNAm in CUD.

Based on epigenome-wide DNAm data, epigenetic clocks provide the opportunity to estimate biological age, a measure that increases naturally with chronological age, but is strongly influenced by disease processes such as chronic inflammation [31]. Two commonly used methodologies are Horvath’s [32] and Levine’s (PhenoAge) [31] epigenetic clocks. Epigenetic age acceleration (EAA), defined as the difference between the estimated epigenetic and chronological age, is of particular interest in the context of diseases, with its interpretation as faster or slower aging considering chronological age as the reference. Existing literature reports on EAA in the PFC of individuals with SUDs such as AUD and OUD compared to a control population, especially for Levine’s clock [29, 33].

In the present study of CUD in human postmortem brain tissue of BA9, we aimed to expand the epigenome-wide characterization of CUD to the prefrontal cortex as another key brain region in addiction. Next to performing an EWAS of CUD in BA9, we aimed to identify functional enrichment of CUD-associated differential methylation within biological pathways and utilized epigenetic clocks to investigate the relationship between chronological and biological age in CUD.

## Materials and Methods Postmortem human brain samples

Postmortem human brain tissue of Brodmann Area 9 from N = 42 male individuals was received from the Douglas Bell Canada Brain Bank (DBCBB, Montréal, Canada). Brain samples were obtained as fresh-frozen tissue and were stored at -80°C until processing. All sample donors were of European ancestry. At the brain bank, phenotypic information was routinely obtained using a psychological autopsy including next-of-kin interviews complemented by the coroner’s notes and the medical record of the deceased donor. Psychiatric diagnoses related to substance use and comorbidities were based on the DSM-IV classification system, however, we used the more recent nomenclature of DSM-5 i.e., substance use disorder instead of substance dependence throughout this study. We included subjects with donor age > 18 and information on CUD status (yes = CUD, no = no CUD). Exclusion criteria were severe neurodevelopmental and neuropsychiatric disorders, SUDs other than CUD or AUD, and psychiatric diagnoses except for depressive disorder diagnoses (MDD, depressive disorder NOS). Matching was performed by age and comorbidities resulting in N =21 CUD case/control pairs.

### DNA extraction and methylation data analysis

Extraction of DNA from N = 42 BA9 postmortem brain samples was performed using the DNeasy Blood & Tissue Kit (Qiagen, Hilden, Germany). DNA was stored at -20°C and epigenome-wide DNA methylation was determined using the Illumina Infinium MethylationEPIC BeadChip (Illumina, San Diego, CA). Samples were randomized based on CUD case/control status and comorbidities such as depressive disorders and AUD prior to the array processing steps.

Quality control (QC) was performed on raw data using a modified version of the CPACOR-pipeline [34] as described in [28]. All statistical analyses were performed using the statistical computing environment R v4.2.1 [35]. In brief, sample exclusion criteria were missing rate > 0.10 and discordance between phenotypic and predicted sex based on DNAm. CpG sites were excluded if 1) the call rate was below 0.95, 2) they are located on sex chromosomes, and 3) their genomic position is related to a genetic variant with MAF > 0.10.

Raw intensities were quantile-normalized followed by a conversion to beta-values of methylation. Beta-values were logit-transformed to M-values as recommended by Du, Zhang [36]. To assess DNAm signatures of CUD in postmortem brain tissue, we applied a linear regression model using M-values as the dependent variable and CUD status as the predictor. Donor age, postmortem interval (PMI), brain pH, diagnosis of a depressive disorder, and alcohol use disorder (AUD) status were included as covariates in the regression model. Based on the methylation data, we estimated the fraction of neuronal and non-neuronal cells with the Houseman approach [37] using the dorsolateral PFC (dlPFC) dataset [38] as reference. We included the fraction of neuronal cells as a covariate to correct for cell-type heterogeneity in the samples. To adjust for technical effects from microarray processing, we derived principal components from the internal control probes of the EPIC array and included the first 10 principal components (cpPC1-cpPC10) into the statistical model.

This resulted in the following linear model:

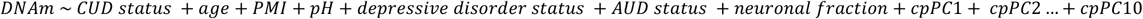

We performed Benjamini-Hochberg (FDR) correction [39] to adjust for multiple testing. Annotation of CpG sites to genes and genomic background was based on the Illumina manifest as downloaded on August 10^th^, 2018: http://webdata.illumina.com.s3-website-us-east-1.amazonaws.com/downloads/productfiles/methylationEPIC/infinium-methylationepic-v-1-0-b4-manifest-file-csv.zip.

To test for enrichment of CUD-associated CpG sites within categories of gene regulatory features (TSS1500, TSS200, 5′-UTR, 1st Exon, ExonBnd, Body, 3′-UTR, and intergenic regions) and the CpG island (CGI) background (N-Shelf, N-Shore, Island, S-Shore, S-Shelf, and open sea regions), we performed chi-squared tests against the distribution of features among all probes on the EPIC array. The results of the enrichment analysis were adjusted for multiple testing using Bonferroni correction.

### Differentially methylated regions

Following the assessment of CUD-associated methylation differences at single CpG sites, we investigated differentially methylated regions associated with CUD status. We used the dmrff algorithm v1.0.0 [40] for the identification of DMRs which was recently shown to outperform several other DMR identification tools when applied to methylation array data [41]. The maximum distance between individual CpG sites (maxgap) was set to 500bp and an association p-value threshold (p.cutoff) of 0.01 was used. The implemented Bonferroni correction was used to adjust for multiple testing during the identification of DMRs.

### Gene-Ontology enrichment analysis

To investigate the potential biological relevance of CUD-associated differential methylation, we analyzed the overrepresentation of differentially methylated CpG sites within biological pathways. Therefore, we applied missMethyl v1.30.0 [42] separately to hypermethylated and hypomethylated CpG sites after selection for a CUD association p-value < 0.001. Correction for multiple testing in missMethyl was performed using FDR.

### WGCNA

To further characterize the biological and functional relevance of CUD-associated methylation signatures in the brain, we performed a weighted correlation network analysis using WGCNA v1.71 [43]. WGCNA identifies co-methylation modules and provides a correlation estimate between modules and traits such as CUD status. We prioritized CpG sites based on their annotation to promoter/transcription start sites (TSS 200 and TSS1500) under the assumption of a cis-regulatory role of their methylation levels on gene expression. Quantile normalized beta values of the resulting 117,876 CpG sites were used as input data for WGCNA. Soft power threshold picking was performed according to the R_signed_^2^ > 0.90 criterion resulting in a power threshold of 5 for the methylation data in BA9. Parameters for constructing the signed network in a block-wise approach were minModuleSize = 30, maxBlockSize = 36,000, and mergeCutHeight = 0.25. For the CUD-associated co-methylation modules, enrichment analysis was performed using missMethyl. Module hub genes were derived by annotation of CpG sites to genes and were defined as the top 0.5% of annotated genes in a co-methylation module ranked by the product of module membership and gene significance. Protein-protein interaction networks of hub genes from the CUD-associated co-methylation modules were generated in STRING v11.5 [44] using an interaction score threshold of 0.7 (high confidence interactions).

### Epigenetic age estimates

Using quantile-normalized beta-values of the BA9 methylation data, we estimated epigenetic age and EAA using epigenetic clocks as implemented in the R package methylclock v1.2.1 [45]. We applied Horvath’s (Horvath) [32] and Levines’s (PhenoAge) [31] epigenetic clocks to assess epigenetic age in the context of CUD directly in the brain. Using a linear regression model, we evaluated the association of CUD status with epigenetic age estimates and EAA while adjusting for covariates such as age, neuronal cell fraction, pH, PMI as well as depressive disorder, and AUD status.

## Results

Demographic data of sample donors are summarized in Table 1, while a detailed overview of toxicology and cause of death is provided in Supplementary Table 1. After quality control of methylation data, all N = 42 subjects, and a total of 654,448 CpG sites remained for further analysis.

**Table 1:** Demographic data of sample donors included in the present study on CUD SD: standard deviation, PMI: postmortem interval, MDD: major depressive disorder, NOS: depressive disorder not otherwise specified, p-value: derived from CUD/no CUD comparison using a t-test for continuous and chi-squared test for categorical variables.

| Characteristic | CUD | No CUD | p-value |
| --- | --- | --- | --- |
| N | 21 | 21 | - |
| Age, years (SD) | 44.4 (13.0) | 50.1 (13.8) | 0.17 |
| Sex (Male/Female) | 21/0 | 21/0 | - |
| pH (SD) | 6.37 (0.39) | 6.26 (0.28) | 0.33 |
| PMI, hours (SD) | 56.8 (23.9) | 58.4 (19.9) | 0.82 |
| Cocaine or cocaine metabolites at death, yes (%) | 15 (71.4) | 0 (0) | - |
| Depressive disorder (MDD, NOS), yes (%) | 9 (42.9) | 5 (23.8) | 0.33 |
| Alcohol use disorder, yes (%) | 9 (42.9) | 4 (19.1) | 0.18 |

### Epigenome-wide association study of CUD

None of the CpG sites passed epigenome-wide significance (p < 7.64e-08). The strongest association with CUD in BA9 was detected for the CpG site cg04659218 (p = 7.98e-07, FDR = 0.52) on chromosome 11. A Manhattan plot summarizing the results is displayed in Figure 1A. Descriptively, several of the CpG sites showing stronger associations with CUD status were annotated to genes encoding transcription factors and DNA binding domain-containing proteins: *ETS Transcription Factor ERG* (*ERG*, p = 4.15e-06), *Zinc Finger And BTB Domain Containing 4* (*ZBTB4*, p = 1.01e-05), *RNA Polymerase II Subunit A* (*POLR2A*, p = 1.01e-05), and *Zinc Finger And BTB Domain Containing 22* (*ZBTB22*, p = 1.97e-05, see Figure 1B). EWAS summary statistics of the 1000 CpG sites displaying the strongest association with CUD status are provided in Supplementary Table 2. A Q-Q plot (λ = 0.969) derived from the EWAS p-values is shown in Supplementary Figure 1.

**Figure 1:**
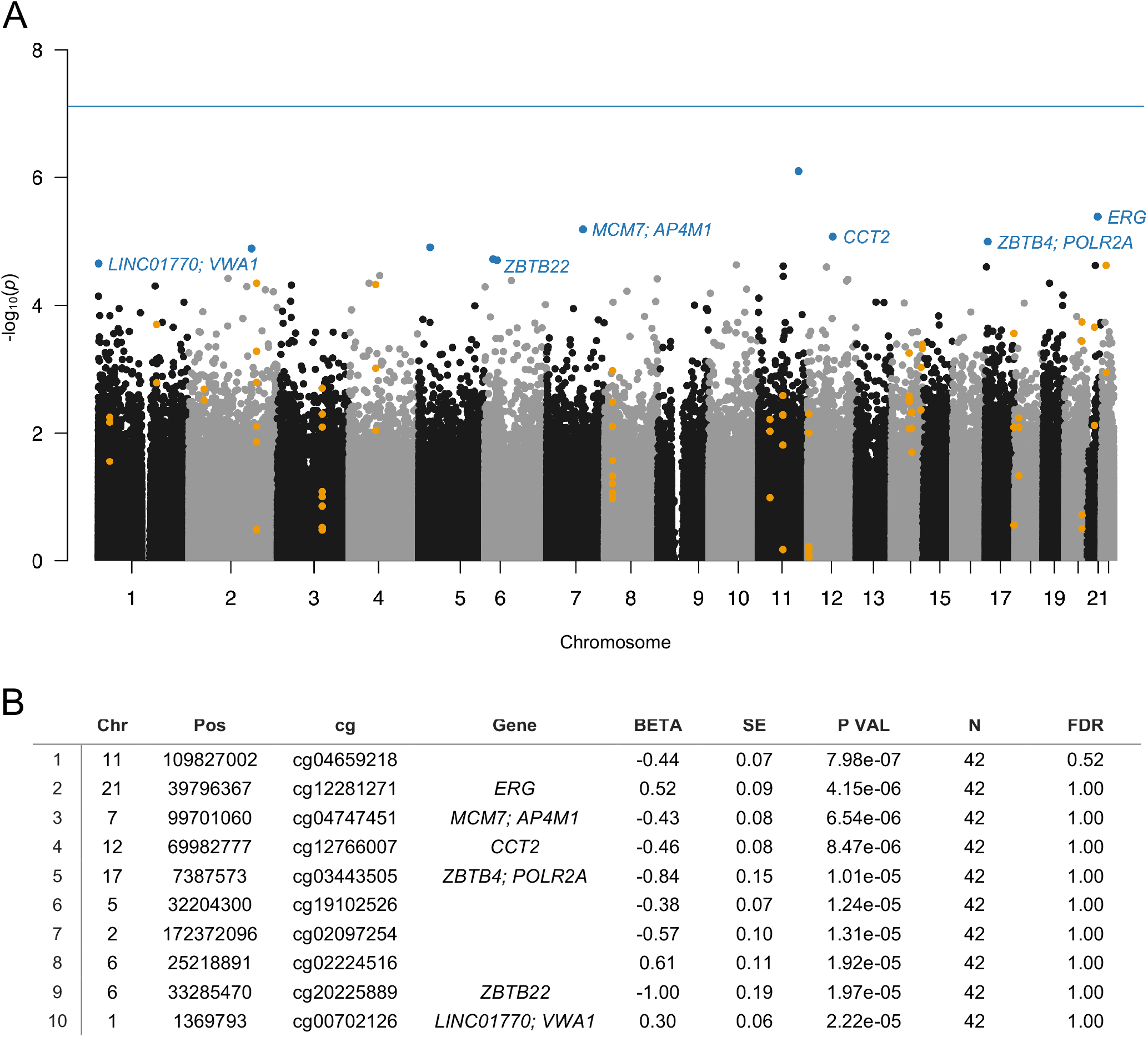
Manhattan plot resulting from the EWAS of cocaine use disorder Results of the EWAS of cocaine use disorder (CUD) assessing N = 654,448 CpG sites in N = 42 postmortem human brain samples from Brodmann Area 9 (BA9). (A) In the Manhattan plot, the 10 CpG sites displaying the strongest association with CUD status were highlighted in blue and were annotated to gene name if available. The 20 CUD-associated DMRs are highlighted in orange. The solid blue line depicts epigenome-wide significance (p < 7.64e-08). (B) EWAS summary statistics for the 10 CpG sites showing the strongest association with CUD status.

### Differentially methylated regions associated with CUD status

A total of 20 DMRs were significantly associated with CUD status after multiple testing correction. We detected more hypermethylated (N = 17, 85%) than hypomethylated (N = 3, 15%) DMRs. The DMR consisting of 6 hypermethylated CpG sites annotated to the genes *LOC101927196* and *Fibrous Sheath Interacting Protein 2 (FSIP2)* displayed the strongest association with CUD status in BA9 (p = 1.74e-16). Results of the DMR analysis including summary statistics for all 20 identified regions are visualized in Table 2.

**Table 2:** CUD-associated differentially methylated regions (N = 20) Differentially methylated regions (DMRs) associated with CUD status. Pos: position of the most significant CpG site within the DMR, BETA, and P VAL: effect size and p-value of this CpG site derived from the EWAS, N CpG: number of CpG sites within DMR, DMR z and DMR q-val: DMR statistics resulting from *dmrff* [40].

|  | Chr | Pos | Gene | BETA | P VAL | DMR start | DMR end | N CpG | DMR z | DMR q-val |
| --- | --- | --- | --- | --- | --- | --- | --- | --- | --- | --- |
| 1 | 2 | 186603233 | *LOC101927196;*<br>*FSIP2* | 0.28 | 4.52e-05 | 186603233 | 186603441 | 6 | 9.71 | 1.74e-16 |
| 2 | 14 | 76734493 |  | 0.63 | 4.69e-03 | 76734327 | 76734550 | 4 | 8.79 | 9.54e-13 |
| 3 | 17 | 79339679 |  | 0.39 | 2.75e-04 | 79339278 | 79339679 | 3 | 7.69 | 9.79e-09 |
| 4 | 11 | 67777715 | *ALDH3B1* | 0.22 | 2.58e-03 | 67777682 | 67777952 | 5 | 7.58 | 2.25e-08 |
| 5 | 4 | 72897782 | *NPFFR2* | 2.69 | 4.74e-05 | 72897613 | 72897782 | 3 | 7.46 | 5.74e-08 |
| 6 | 18 | 11751352 | *GNAL* | 0.24 | 5.76e-03 | 11751317 | 11751352 | 3 | 6.99 | 1.84e-06 |
| 7 | 3 | 123813596 | *KALRN* | 0.33 | 1.98e-03 | 123813383 | 123813596 | 8 | 6.95 | 2.40e-06 |
| 8 | 8 | 22132992 | *LOC100507071;*<br>*PIWIL2* | 0.72 | 1.05e-03 | 22132641 | 22132992 | 8 | 6.62 | 2.38e-05 |
| 9 | 21 | 30449989 | *MAP3K7CL* | 0.47 | 2.20e-04 | 30449989 | 30450130 | 2 | 6.50 | 5.30e-05 |
| 10 | 22 | 30476204 | *HORMAD2-AS1;*<br>*HORMAD2* | 0.64 | 2.38e-05 | 30476204 | 30476206 | 2 | 6.35 | 1.47e-04 |
| 11 | 1 | 161008297 | *TSTD1* | 0.41 | 1.99e-04 | 161008127 | 161008297 | 2 | 6.32 | 1.68e-04 |
| 12 | 20 | 47896888 | *ZFAS1;*<br>*SNORD12B;*<br>*SNORD12* | 0.34 | 1.83e-04 | 47896888 | 47897207 | 4 | 5.97 | 1.59e-03 |
| 13 | 2 | 42274082 | *PKDCC* | 0.42 | 2.05e-03 | 42274082 | 42274306 | 2 | 5.89 | 2.62e-03 |
| 14 | 12 | 4382184 | *CCND2-AS1;*<br>*CCND2* | -0.39 | 5.01e-03 | 4382114 | 4382184 | 6 | -5.81 | 4.07e-03 |
| 15 | 14 | 105915268 | *MTA1* | 0.25 | 4.08e-04 | 105915004 | 105915268 | 2 | 5.74 | 6.17e-03 |
| 16 | 14 | 101908830 | *LOC100507277* | -0.33 | 9.41e-04 | 101908830 | 101908865 | 2 | -5.66 | 1.02e-02 |
| 17 | 14 | 70193496 |  | -0.41 | 2.63e-03 | 70193468 | 70193496 | 3 | -5.58 | 1.56e-02 |
| 18 | 14 | 69656853 | *EXD2* | 0.45 | 5.58e-04 | 69656853 | 69657279 | 2 | 5.58 | 1.62e-02 |
| 19 | 11 | 32421635 | *WT1* | 0.34 | 6.09e-03 | 32421514 | 32421635 | 3 | 5.41 | 4.28e-02 |
| 20 | 1 | 32169770 | *COL16A1* | 0.35 | 5.64e-03 | 32169701 | 32169770 | 3 | 5.38 | 4.89e-02 |

### Functional annotation and enrichment analysis of CUD-associated CpG sites

To analyze enrichment within genomic features (Figure 2A) and the CGI background (Figure 2B), we prioritized our findings based on CpG sites associated with CUD at nominal significance (p < 0.05). This resulted in a subset of N = 29,176 CpG sites of which N = 14,737 were hypermethylated and N = 14,439 were hypomethylated. We detected a balanced ratio between hyper- and hypomethylation in this subset (Figure 2C). For the CUD-associated CpG sites, a statistically significant enrichment within 1st Exon, N-Shore, and CpG island regions was detected, whereas a significant depletion from intergenic and open sea regions was found.

**Figure 2:**
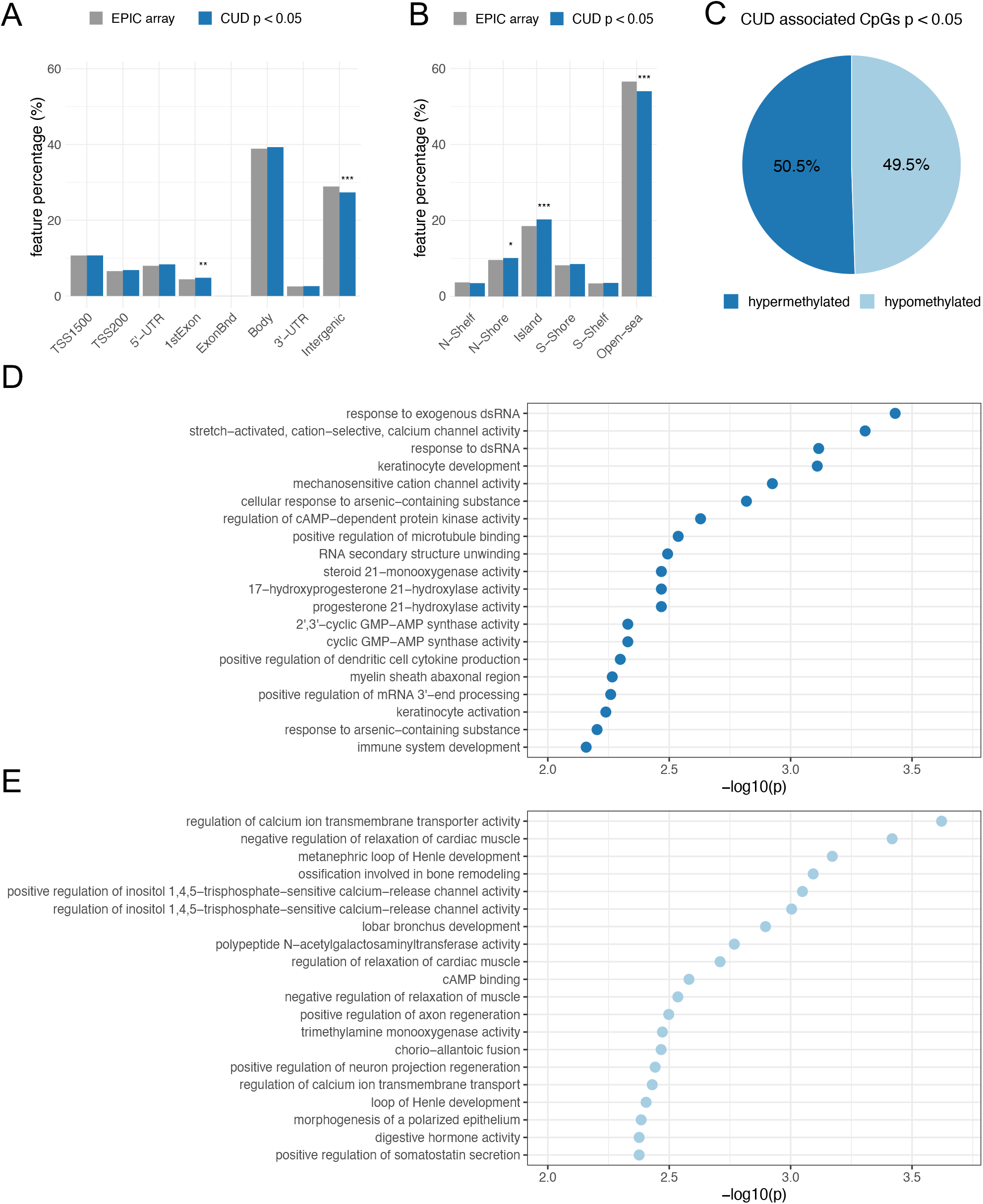
Genomic feature annotation and GO enrichment analysis of CUD-associated CpG sites in BA9 Results from the EWAS of CUD were prioritized based on association p-values resulting in a subset of N = 29,176 CpG sites with p_assoc_ < 0.05 that was further analyzed by annotation to (A) gene regulatory features and (B) the CpG island background. The proportion of CUD-associated CpG sites (blue) within different groups was compared to the EPIC array background (gray). Differences remaining statistically significant after Bonferroni multiple testing correction are highlighted using asterisks (* = p_adj_ < 0.05, ** = p_adj_ < 0.01, *** = p_adj_ < 0.001). (C) Hypermethylation (light-blue) and hypermethylation (dark blue) among the N = 29,176 CUD-associated CpG sites. The top 20 CUD-associated GO terms resulting from missMethyl [42] are shown for (D) the N = 14,737 hypermethylated and (E) the N = 14,439 hypomethylated CpG sites.

The most significantly enriched GO term was *response to exogenous dsRNA* (p = 3.71e-04) for hypermethylated CpG sites and *regulation of calcium ion transmembrane transporter activity* (p = 2.39e-04) for the subset of hypomethylated CpG sites. No enrichment remained statistically significant after multiple testing correction (Supplementary Table 3). Descriptively, genes harboring hypermethylated CpG sites were overrepresented within biological pathways involved in cation and especially mechanosensitive calcium ion channel activity, cAMP-dependent protein kinase (PKA) activity, and immunological signaling mainly related to the CGAS-mediated response to nucleic acids (Figure 2D). In contrast, the gene set derived from hypomethylated CpG sites was enriched within pathways related to calcium transmembrane transport, negative regulation of muscle relaxation, and cAMP binding (Figure 2E). Also, positive regulation of inositol-1,4,5-trisphosphate-sensitive calcium release channel activity was among the enriched GO terms detected for hypomethylated CpG sites.

### Weighted correlation network analysis

Due to the known cis-regulatory role of DNAm on gene expression, we extracted TSS1500- and TSS200-associated CpG sites from the methylation matrix to identify transcriptionally relevant co-methylation networks. After hierarchical clustering in WGCNA, the sample tree was pruned resulting in the removal of one sample from the analysis (N = 41 remaining). The sample dendrogram was related to traits resulting in the clustering of CUD cases and non-CUD individuals within subgroups (Figure 3A). Network construction led to the identification of a total of 69 modules ranging in size between 38 and 35,758 CpG sites. A module-trait correlation heatmap for all resulting modules is shown in Supplementary Figure 2.

**Figure 3:**
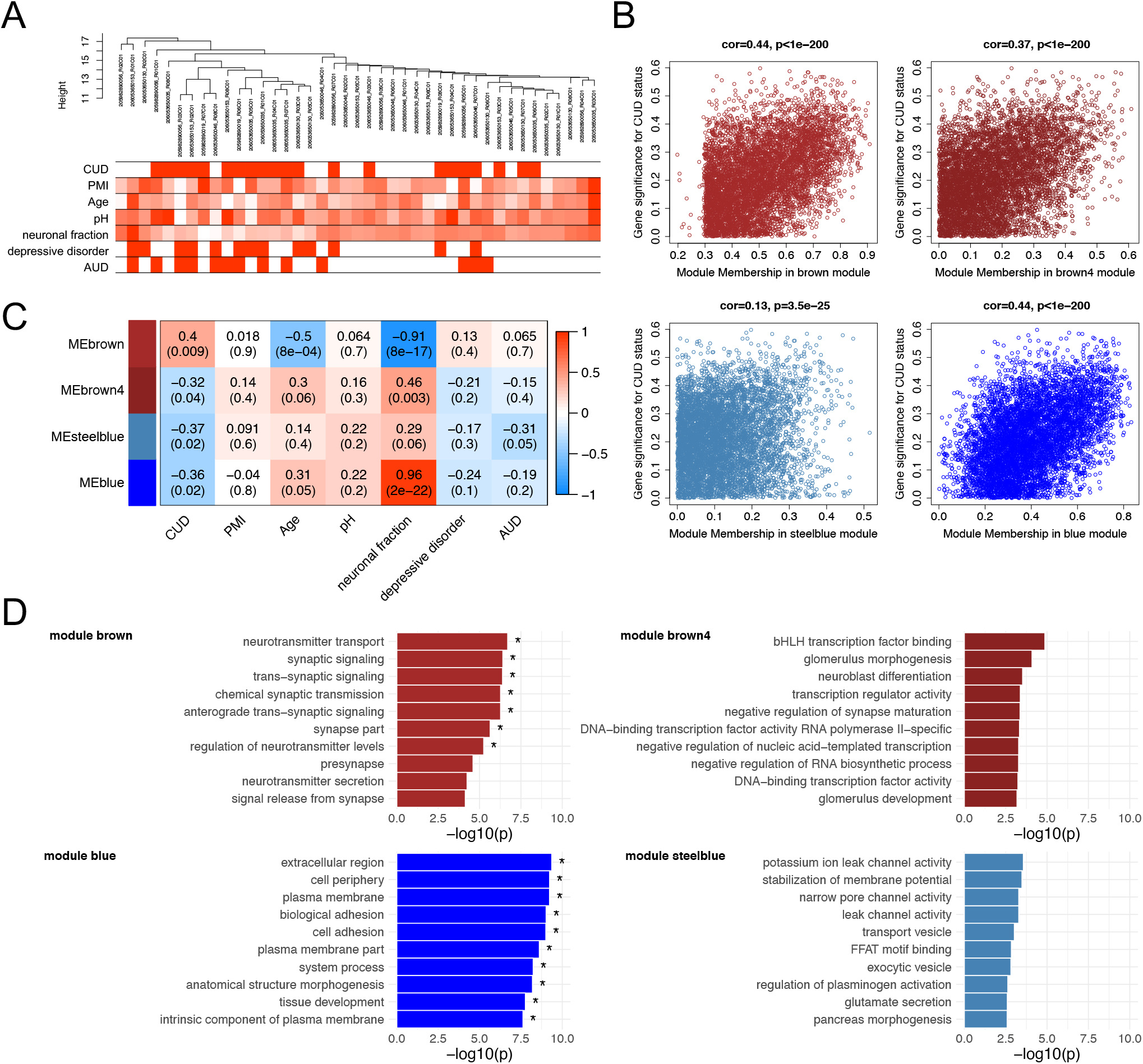
Functional analysis of the CUD-associated co-methylation modules identified in WGCNA Quantile-normalized beta values of methylation for N = 117,876 promoter-associated CpG sites (TSS200 and TSS1500) were extracted and used as an input dataset for WGCNA. (A) Dendrogram of hierarchical sample clustering with trait annotation. (B) Correlation plots of module membership and CUD gene significance for the 4 CUD-associated co-methylation modules (brown, brown4, steelblue, blue). (C) Module-trait relationships for the CUD-associated modules. Module-trait correlation coefficients are color-coded, and correlation p-values are shown in brackets. (D) Results of the GO enrichment analysis. The 10 GO terms with the strongest enrichment within the respective co-methylation module are displayed (* = FDR < 0.05). CUD: cocaine use disorder, PMI: postmortem interval, neuronal fraction: estimated neuronal cell type proportion based on Houseman et al. [37], AUD: alcohol use disorder.

Of the 69 modules, 4 were significantly associated with CUD status: brown (N = 6,309 CpG sites, p = 9.15e-03), brown4 (N = 187 CpG sites, p = 0.04), blue (N = 14,050 CpG sites, p = 0.02), and steelblue (N = 340 CpG sites, p = 0.02). A positive, highly significant correlation between module membership and gene significance for CUD status was detected within these modules (Figure 3B). Whereas module brown was positively associated with CUD status (r = 0.40), a negative correlation was identified for modules brown4, blue, and steelblue (-0.37 ≤ r ≤ -0.32, see Figure 3C). In addition, for the brown module, significant negative correlations were detected with age (r = -0.5, p = 8.11e-04) and the fraction of neuronal cells (r = -0.91, p = 7.69e-17). The blue module also depicts a cell type-specific network displaying a highly significant positive correlation with neuronal fraction (r = 0.96, p = 1.78e-22). To further assess the biological processes related to CUD-associated modules, we performed GO enrichment analyses (Figure 3D, Supplementary Table 4). After multiple testing correction, GO enrichment in modules brown and blue remained statistically significant. Descriptively, module brown was enriched for synaptic signaling and neurotransmitter transport, whereas module brown4 revealed GO terms related to transcription factor binding and neuroblast as well as synapse maturation. The blue module displayed enrichment of large GO terms involved in basic cellular processes and compartments, such as extracellular region, plasma membrane, and cell adhesion. Within module steelblue, nominally significant pathway enrichment was related to ion channel activity, membrane potential homeostasis, and vesicle transport. Full enrichment statistics for the top enriched GO terms in each module are provided in Supplementary Table 5.

To further characterize the molecular processes enriched in the CUD-associated modules, we constructed protein-protein interaction (PPI) networks based on module hub genes (for network plots see Supplementary Figure 3). Among PPI network hub nodes of modules brown and blue (Supplementary Table 6), we found members of the TGF-β signaling cascade such as *Transforming Growth Factor Alpha* (*TGFA*), *Transforming Growth Factor Beta 1* (*TGFB1*), and *Endoglin* (*ENG*). Reflecting the pathway enrichment for neuron-related biological processes in modules brown, brown4, and steelblue, we detected multiple hub nodes with an important role in neuronal activity and function within these modules. Among them were the Ca_v_1.2 subunit *Calcium Voltage-Gated Channel Subunit Alpha1 C* (*CACNA1C*, module steelblue), the glucocorticoid receptor *Nuclear Receptor Subfamily 3 Group C Member 1* (*NR3C1*, module brown), the acetylcholine transporter *Solute Carrier Family 18 Member A3* (*SLC18A3*, module brown4), and *Vesicle Associated Membrane Protein 2* (*VAMP2*, module steelblue). Further, several tyrosine kinases and their downstream targets were found among hub nodes of PPI networks derived from modules brown4 and blue, such as *Fibroblast Growth Factor Receptor 1* (*FGFR1*), *Erb-B2 Receptor Tyrosine Kinase 3* (*ERBB3*), *KRAS Proto-Oncogene GTPase* (*KRAS*), and *Signal Transducer And Activator Of Transcription 5A* (*STAT5A*). The *Jun Proto-Oncogene, AP-1 Transcription Factor Subunit* (*JUN*) was the node displaying the strongest connectivity in module steelblue.

### Epigenetic age in CUD

A moderate (R_Levine_^2^ = 0.44) to strong (R_Horvath_^2^ = 0.80) correlation was found between estimated epigenetic age and chronological age (Figure 4A and 4B). When assessing EAA using Horvath’s and Levine’s epigenetic clocks while adjusting for covariates, we observed a trend toward a more positive EAA in CUD cases compared to individuals without CUD, however, not statistically significant (p_Horvath_ = 0.392, p_Levine_ = 5.12e-02, Figure 4C, and 4D). For both epigenetic clocks, we consistently found positive associations for CUD status in the regression models for epigenetic age and EAA (Supplementary Table 7).

**Figure 4:**
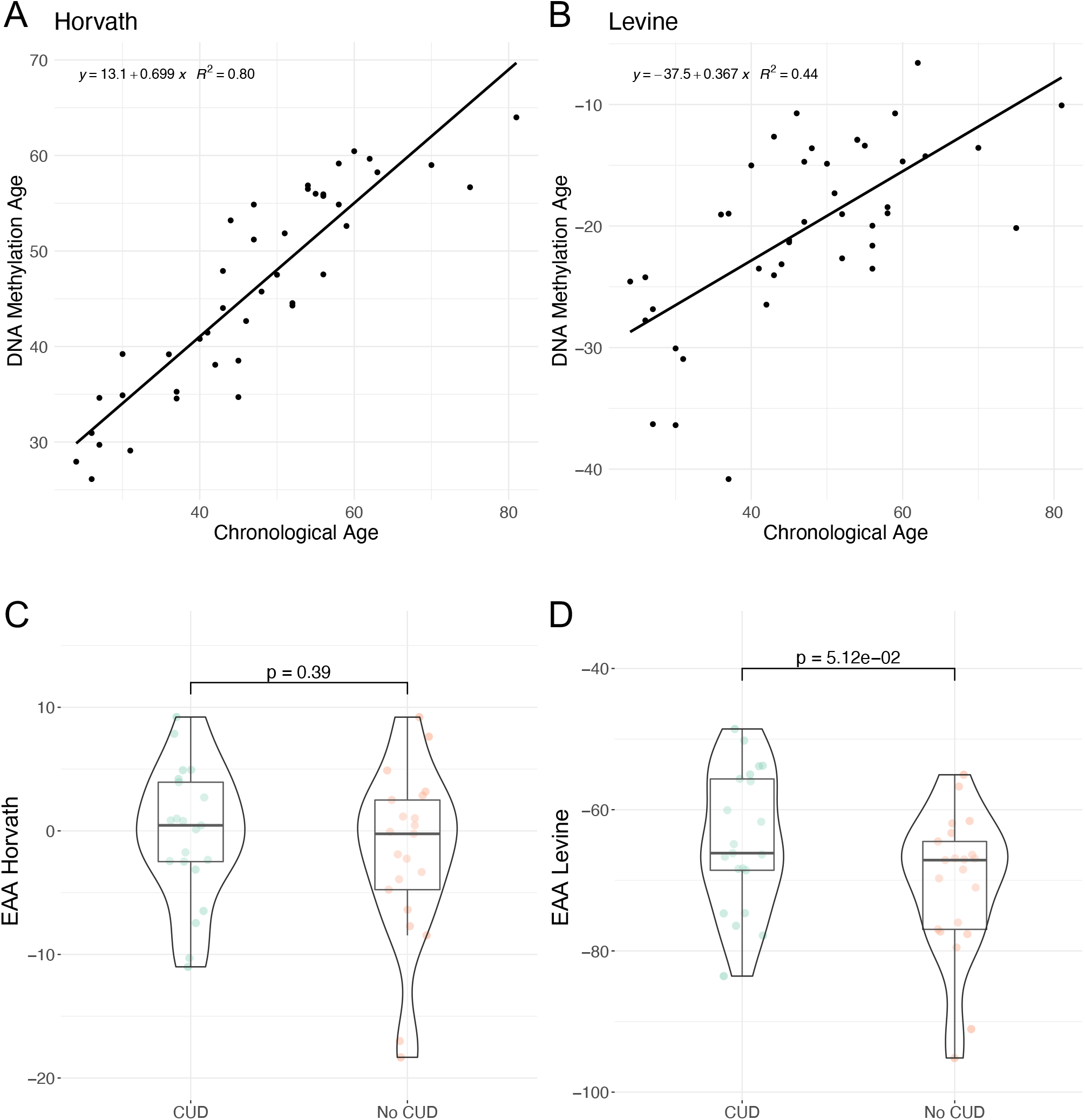
Evaluation of epigenetic clocks in CUD using postmortem human brain samples of Brodmann Area 9 Correlation of chronological age with DNA methylation based (A) Horvath’s and (B) Levine’s epigenetic age estimates as implemented in the R package methylclock [45]. Comparison of epigenetic age acceleration (EAA) between individuals with and without CUD using (C) Horvath’s and (D) Levine’s epigenetic clocks.

## Discussion

We performed an epigenome-wide analysis of DNAm differences in BA9 postmortem human brain tissue from N = 21 CUD cases and N = 21 individuals without CUD. In the BA9 subregion of the human prefrontal cortex, none of the associations of CpG site methylation levels with CUD status passed the level of epigenome-wide significance. However, we identified 20 DMRs significantly associated with CUD.

Among the strongest single-site associations with CUD status, there were multiple CpG sites annotated to genes involved in transcriptional regulation either via intrinsic transcription factor activity or due to the interaction with RNA polymerase II (POL-II). The transcription factor ERG is involved in the regulation of vasculature integrity [46]. For ZBTB4, binding to methylated CpG sites was reported [47] and ZBTB22 is assumed to be involved in transcription regulation via interaction with POL-II [48]. In our EWAS, we also detected CUD-associated differential methylation within the largest subunit of POL-II (POL2RA), suggesting a role of CUD-associated methylation signatures in gene expression regulation. Results of the enrichment analysis for CUD-associated CpG sites within genomic features are in line with this: the significant enrichment of CUD-associated differential methylation within features containing transcription factor binding sites and the depletion from intergenic and non-CGI-associated (open sea) regions points towards a regulatory potential of CUD-associated DNAm patterns on gene expression.

For several of the genes harboring DMRs significantly associated with CUD in BA9, there is evidence for their relevance for neuronal function. The DMR displaying the strongest association with CUD status was related to the *LOC101927196* gene. Overexpression of the long non-coding RNA LOC101927196 was shown to reduce cell proliferation and support apoptotic processes in the brain in a rat model of autism [49]. For *NPFFR2* and *KALRN*, we found direct links to their importance for the biological response to cocaine. Neuropeptide FF Receptor 2 (NPFFR2) interferes with the opioid system in the brain and its activation contributes to anxiety-like behavior in mice [50]. Intraventricular application of NPFF reduced cocaine-induced conditioned place preference in rats pointing toward an important role of NPFF signaling in cocaine reward processing [51]. The *Kalirin RhoGEF Kinase (KALRN*) gene encodes several isoforms and Kalirin-7 was shown to be involved in the development and function of dendritic spines [52]. After repeated cocaine exposure in a mouse model, expression levels of Kalirin-7 were elevated in the NAc contributing to the formation of new dendritic spines. Kalirin-7 knock-out mice displayed increased cocaine-induced locomotor sensitization and a decreased conditioned place preference [53].

Looking at multiple brain regions allows a more complete understanding of the role of DNAm alterations in CUD. In line with results from previous studies on epigenome-wide alterations of CUD in the CN [23] and in the NAc [24], we observed more hypermethylation than hypomethylation at the DMR level in BA9. We detected one DMR annotated to the *Aldehyde Dehydrogenase 3 Family Member B1* (*ALDH3B1*) gene on chromosome 11 that was identified in both, our analysis in BA9 and in the previous study in the CN. Hypermethylation of this DMR was also consistent between studies. It has to be noted that the EPIC array covers only a finite number of CpG sites which may limit the comparison with results from reduced representation bisulfite sequencing.

No enrichment of hypo- and hypermethylated CUD-associated CpG sites among GO terms remained statistically significant after multiple testing correction. The functional relevance of CUD-associated differential methylation should thus be evaluated together with the enriched biological processes and hub genes derived from WGCNA co-methylation modules. Three of the four CUD-associated modules were enriched for neuron-specific processes such as synaptic signaling, ion channel activity, and neurotransmitter transport. Next to the association with CUD, modules brown and blue displayed a strong correlation with neuronal fraction, which might also contribute to the enrichment of neuronal processes within these co-methylation modules. PPI networks based on module hub genes revealed *CACNA1C* and *NR3C1* as highly connected genes for which a previous relation to cocaine use is known. *CACNA1C* is a well-described risk gene for psychiatric disorders such as bipolar disorder, MDD, and schizophrenia [54]. By encoding a subunit of the voltage-gated Ca_v_1.2 calcium channel, it depicts a key regulator of neuronal excitability. Further, the expression of cocaine-induced locomotor sensitization was shown to be dependent on Ca_v_1.2 signaling in mice [55]. The *NR3C1* gene encodes a glucocorticoid receptor for which downregulation in blood was observed in chronic cocaine users [56] and its DNAm levels were shown to predict substance use phenotypes in adolescents [57]. Further, brain-specific conditional knock-out of *NR3C1* flattened the dose-response curve for cocaine self-administration and reduced behavioral sensitization in an intravenous cocaine self-administration model in mice [58]. Thus, *CACNA1C* and *NR3C1* depict important regulators of the reinforcing effects of cocaine.

We further identified *JUN* as the hub node with the strongest connectivity in module steelblue. By forming heterodimers with FOS family proteins, JUN is involved in the formation of the AP-1 transcription factor complex depicting a key regulator of neuroplasticity in addiction [59]. In a mouse model, decreased cocaine-induced conditioned place preference was found during expression of a dominant-negative mutant of c-Jun in the brain [60]. Epigenetic dysregulation of the JUN interaction network might therefore contribute to neurocircuit alterations in CUD.

In our analysis of epigenetic age in the brain, we observed a trend towards an EAA in CUD cases compared to individuals without CUD that was more pronounced with Levine’s clock compared to Horvath’s clock. This is in line with results from previous studies that report on EAA, particularly for Levine’s epigenetic clock, in the PFC of individuals with opioid intoxication, AUD, and OUD [29, 33]. As cocaine intake is associated with neuroinflammatory processes in the brain [61, 62], EAA in CUD could be a consequence of an inflammatory response that is particularly reflected by Levine’s epigenetic clock [31]. However, as comorbid diseases are frequently observed in CUD, an increase in EAA could at least in part be driven by systemic diseases that are themselves associated with an accelerated epigenetic age. Due to the lack of information on somatic comorbidities, we were not able to address this point in the present study. Further, for the interpretation of epigenetic age in the brain, it must be noted that epigenetic clocks are at least to some extent tissue-specific and might not be applicable to every tissue. We exclusively assessed epigenetic clocks that were either trained also on non-blood tissues (Horvath) or at least showed a high correlation of findings across tissues (Levine). The strong negative values of Levine’s EAA we observed in all brain samples might represent a postmortem brain tissue artifact, as Levine’s clock was initially trained on blood methylation data. Analyses with larger sample sizes, careful adjustment for comorbidities, and appropriate estimation methods are required to evaluate if EAA in the brain is a true phenomenon in CUD.

Several limitations apply to our study on DNAm signatures of CUD in postmortem human brain tissue. First, our sample size of N = 42 is comparatively low for an epigenome-wide analysis and is associated with limitations in statistical power. Another limitation is the frequent comorbidity of CUD with other SUDs and mood disorders, which is well reflected in our sample. Drug intoxication at death and medication prior to death could have affected methylation levels and therefore might confound the results for CUD. Next to the CUD phenotype, assessment of used substances and comorbidities including their treatment is crucial to adjust for confounding effects on DNAm signatures. Due to the small sample size, we were not able to adjust for drug intoxication at death and medication prior to death. Thus, we cannot rule out a potential confounding of the identified CUD-associated methylation signatures by epigenetic effects of other substances. Third, the case-control design of the EWAS does not allow the assessment of temporal changes in DNAm levels. Thus, within the present study, we are not able to separate differential methylation acquired during the disease course of CUD from an epigenetic predisposition to CUD.

In the present study, we identified CUD-associated DNAm signatures in the BA9 subregion of the human PFC. We detected CUD-associated differential methylation within several transcription factor genes, an enrichment within genomic features of transcription regulation, and an association with gene regulatory networks involved in neurotransmission and neuroplasticity. Specifically, epigenetic alterations related to calcium and cAMP signaling as well as the *CACNA1C* and *JUN* PPI networks might contribute to the development of the altered neurocircuits observed in the CUD brain.

To follow up on these hypotheses, further studies are required. These should be based on larger sample sizes with detailed phenotyping to minimize the confounding of results in association studies. Further, integration of epigenome-wide methylation data with other omics data such as transcriptomics or proteomics is necessary to prioritize and substantiate findings from the EWAS. Such multi-omics studies depict promising approaches for deeper profiling of altered biological processes in SUDs as recently shown for AUD [63] and opioid use disorder [64]. To address the temporal dynamics of molecular changes in the disease course of CUD, a longitudinal study assessing different stages of CUD within the same individuals should be performed. While a longitudinal design is challenging using postmortem brain tissue, it depicts a feasible approach for blood samples. The application of such innovative methods could pave the way for the development of precision medicine approaches thereby addressing the urgent need for novel therapeutic options in CUD.

## Data Availability

Raw data and full summary statistics are available upon request.

## Ethics approval statement

Postmortem brain tissue sampling at the Douglas Bell Canada Brain Bank was performed according to their established ethical standards including written informed consent from the next-of-kin for each subject. Our study design was approved by the Ethics Committee II of the University of Heidelberg, Medical Faculty Mannheim, Germany, under the register number 2021-681.

## Author Contributions

Conceptualization, S.H.W., R.S., A.C.H., and M.R.; data curation, E.P., F.S., J.C.F., and L.Z.; formal analysis, E.P., and L.Z.; funding acquisition, S.H.W., R.S., A.C.H., and M.R.; investigation, E.P., L.Z., J.C.F., M.M.F., A.C.H., R.S., and S.H.W.; methodology, L.Z., E.P., F.S., and J.F.; project administration, S.H.W., M.M.N., A.C.H., and R.S.; resources, G.T., N.M., A.C.H., and M.M.N.; supervision, S.H.W, J.F., M.R., and R.S.; writing—original draft, E.P., L.Z.; writing—review and editing, E.P., L.Z., F.S., J.F., J.C.F., M.M.F., A.C.H., G.T., N.M., M.M.N., R.S., M.R., and S.H.W.

## Acknowledgments

We thank Claudia Schäfer-Arnold and Elisabeth Röbel for technical assistance.

## Data Availability Statement

Raw data and full summary statistics are available upon request.

