## Supplementary Figures for "DNA Methylation in Cocaine Use Disorder – An Epigenome-wide Approach in the Human Prefrontal Cortex"

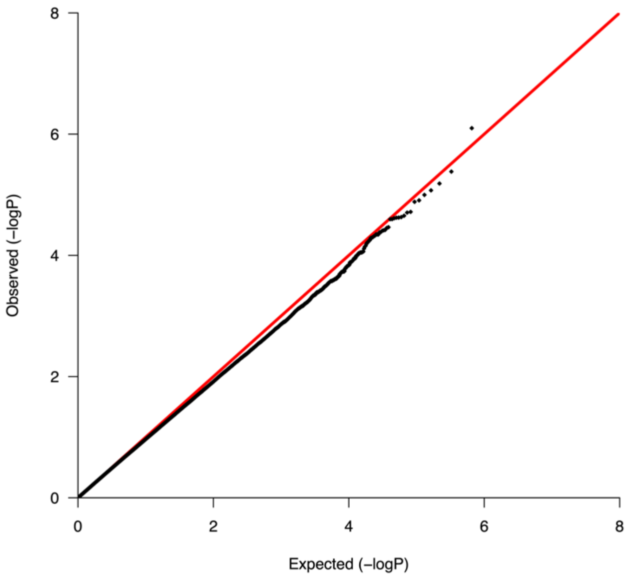


Supplementary Figure 1: QQ-Plot (λ = 0.969) of the EWAS of cocaine use disorder in human postmortem brain tissue of Brodmann Area 9 (N = 42).

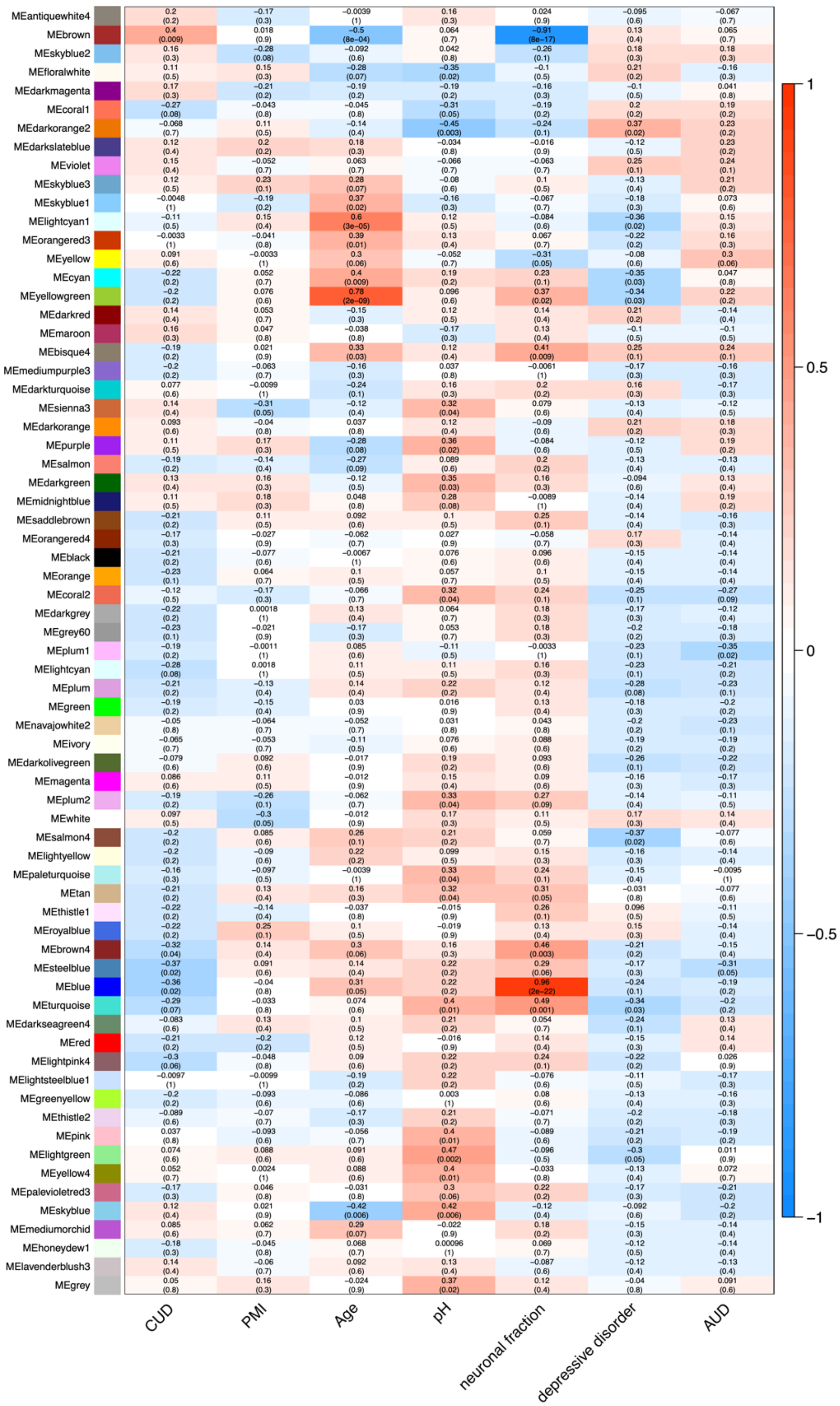


Supplementary Figure 2: Module-trait correlation plot displaying all co-methylation modules resulting from WGCNA.

CUD: cocaine use disorder, PMI: postmortem interval, neuronal fraction: estimated neuronal cell type proportion, AUD: alcohol use disorder.

A module brown

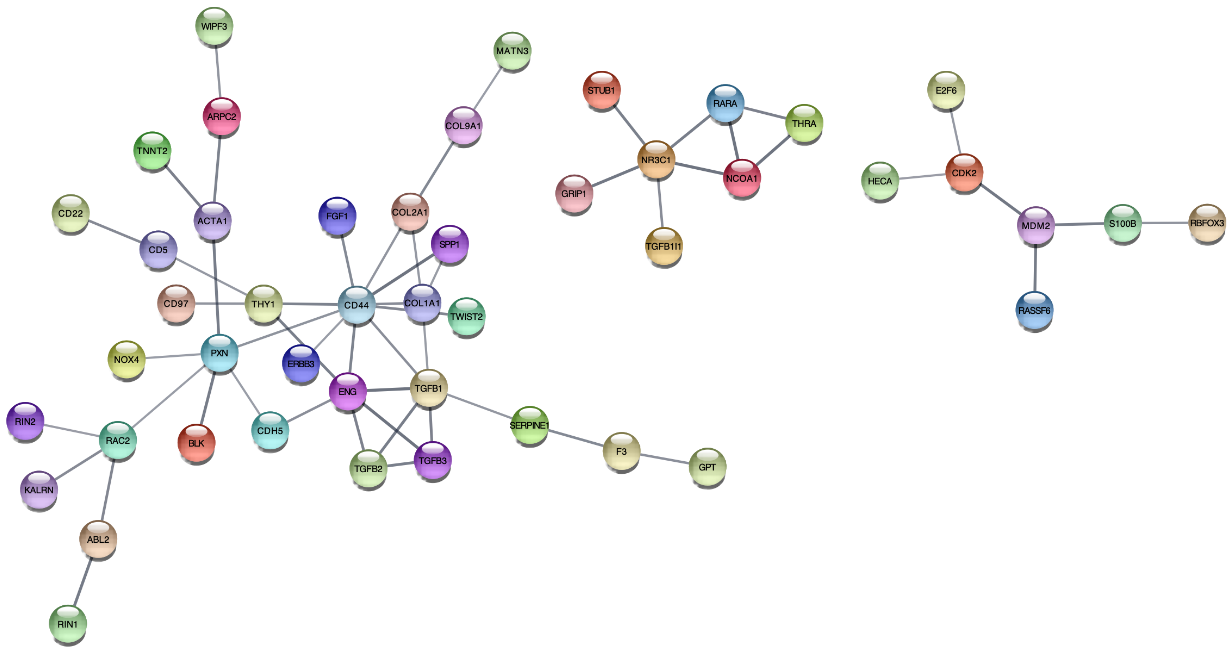


B module brown4

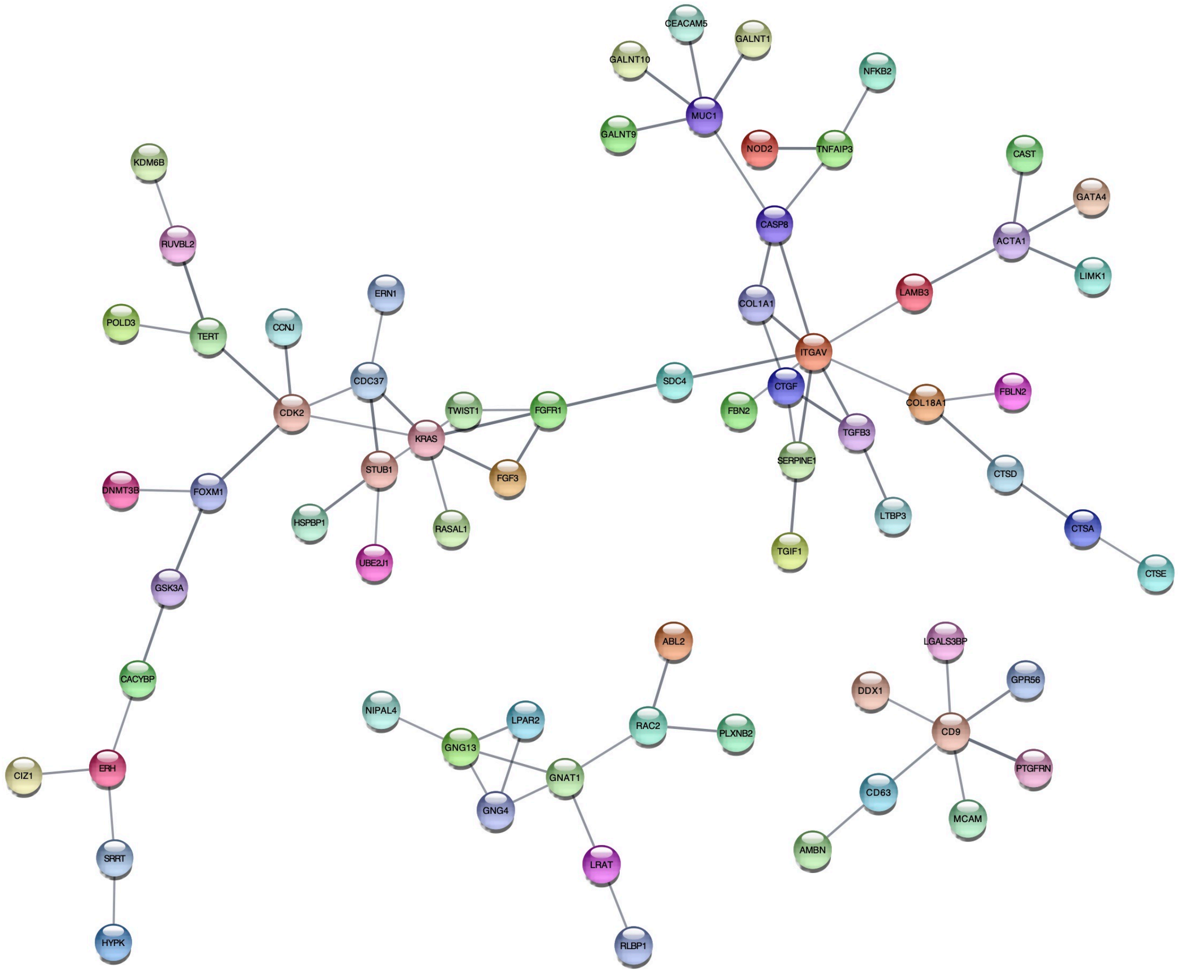

C module blue

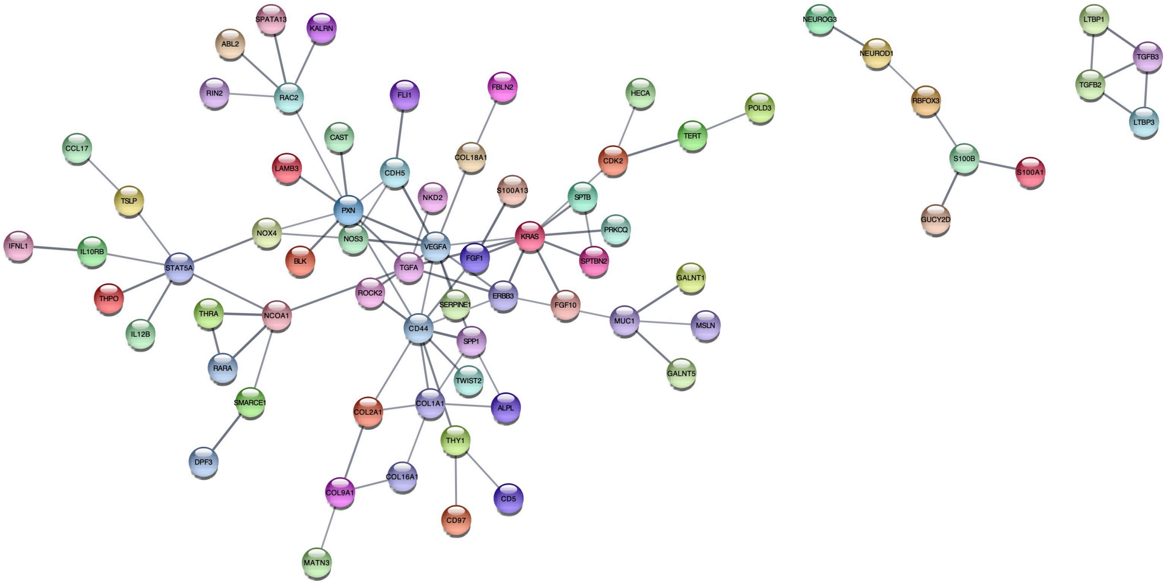


D module steelblue

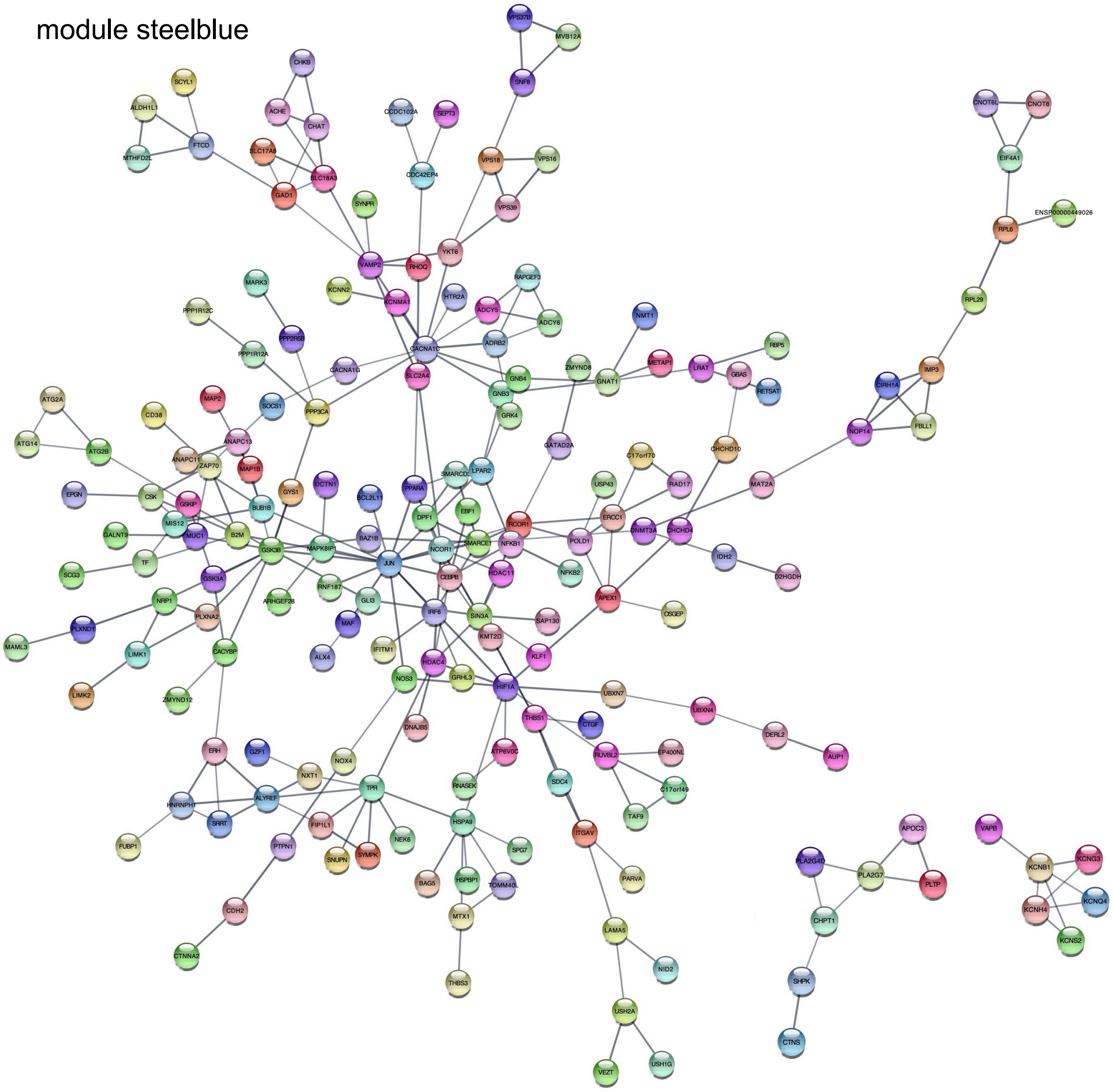

Supplementary Figure 3: Protein-protein-interaction networks derived from CUD-associated co-methylation module hub genes.

The top three networks ranked by size and the connectivity of nodes are shown for WGCNA-derived co-methylation modules (A) brown, (B) brown4, (C) blue, and (D) steelblue. Network plots were generated using the *Search Tool for the Retrieval of Interacting Genes/Proteins* (STRING, v.11.5) with an interaction score threshold of 0.7 (high confidence interactions).
